# Duration of viable virus shedding in SARS-CoV-2 omicron variant infection

**DOI:** 10.1101/2022.03.01.22271582

**Authors:** Julie Boucau, Caitlin Marino, James Regan, Rockib Uddin, Manish C. Choudhary, James P. Flynn, Geoffrey Chen, Ashley M. Stuckwisch, Josh Mathews, May Y. Liew, Arshdeep Singh, Taryn Lipiner, Autumn Kittilson, Meghan Melberg, Yijia Li, Rebecca F. Gilbert, Zahra Reynolds, Surabhi L. Iyer, Grace C. Chamberlin, Tammy D. Vyas, Marcia B. Goldberg, Jatin M. Vyas, Jonathan Z. Li, Jacob E. Lemieux, Mark J. Siedner, Amy K. Barczak

**Affiliations:** Ragon Institute of MGH, MIT and Harvard, Cambridge, MA, USA; Brigham and Women’s Hospital Boston, MA, USA; Massachusetts General Hospital, Boston, MA, USA; Harvard Medical School, Boston, MA, USA; Broad Institute, Cambridge, MA, USA

## Abstract

Clinical features of SARS-CoV-2 Omicron variant infection, including incubation period and transmission rates, distinguish this variant from preceding variants. However, whether the duration of shedding of viable virus differs between omicron and previous variants is not well understood. To characterize how variant and vaccination status impact shedding of viable virus, we serially sampled symptomatic outpatients newly diagnosed with COVID-19. Anterior nasal swabs were tested for viral load, sequencing, and viral culture. Time to PCR conversion was similar between individuals infected with the Delta and the Omicron variant. Time to culture conversion was also similar, with a median time to culture conversion of 6 days (interquartile range 4-8 days) in both groups. There were also no differences in time to PCR or culture conversion by vaccination status.

## MAIN

The omicron variant of SARS-CoV-2 has a shorter incubation period and substantially higher transmission rates than prior variants, dwarfing preceding variants in globally reported cases^1-3^. Recently, public health guidance has recommended shortening the strict isolation period in non-health care settings from 10 to 5 days after symptom onset or the initial positive test^4^. However, viral decay kinetics and duration of shedding viable virus for the omicron variant have not been well characterized.

We followed symptomatic outpatients newly diagnosed with COVID-19 with longitudinal sampling of nasal swabs for viral load, sequencing, and viral culture^5^. A subset of specimens also underwent laboratory-based antigen testing. During July 2021 – January 2022 we enrolled 56 individuals, including 37 sequenced as delta and 19 sequenced as omicron variant infections. All but one participant had symptomatic infection. Participant characteristics were similar between groups, with the exception of a higher vaccine boosting rate among those with omicron infection (26% vs 5%, Table 1). Viral load decay and time to negative PCR did not differ between participants infected with omicron vs. delta (adjusted hazard ratio [AHR] 0.85, 95%CI 0.44, 1.61, Figure 1A-B, Table 2). Duration of shedding of viable virus, as measured by time to culture conversion, was also similar by variant (AHR 0.86, 95%CI 0.47, 1.58, Figure 1C, Table 3), with a median time to culture conversion of 6 days in both groups (IQR 4-8 days, Figure 1C). In the overall cohort, there were no differences in time to PCR conversion (*P*>0.08) or culture conversion (*P*>0.57) by vaccination status (Figures 1D-E, Tables S2 and S3). Laboratory-based antigen testing of specimens stored in viral transport media had a specificity of 88% (95%CI, 71-96%) for culture positivity between days 6-10 after infection (Figure 2, Table 4).

**Table 1.** Cohort characteristics

|  | Omicron variant<br>infection (n=19) | Delta variant<br>infection (n=37) | P-value |
| --- | --- | --- | --- |
| Female, n (%) | 14 (74%) | 23 (62%) | 0.55 |
| Age, mean (SD) | 39 (14) | 42 (16) | 0.61 |
| Vaccination status, n (%) |  |  | 0.04 |
| Unvaccinated | 1 (5%) | 9 (24%) |  |
| Vaccinated | 13 (68%) | 26 (70%) |  |
| Boosted | 5 (26%) | 2 (5%) |  |
| Days since last vaccination, mean (SD) | 169 (130) | 192 (77) | 0.88 |
| Symptomatic infection, n (%) | 19 (100%) | 36 (97%) | >0.99 |

**Figure 1.**
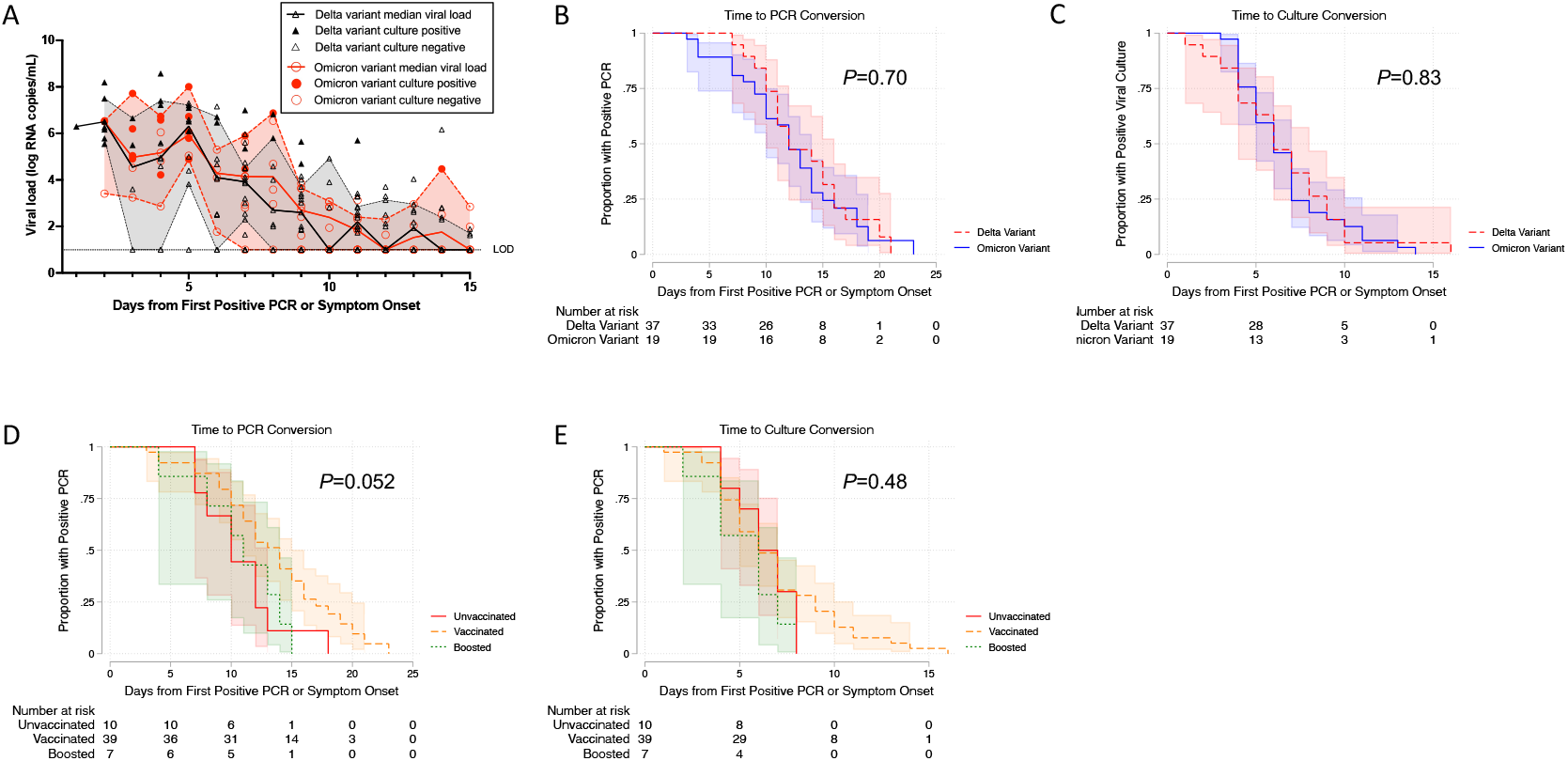
Virologic decay from time of first positive PCR or symptom onset. Observations indicate viral loads from nasal swabs from individual patient samples. Fit indicates the median viral load at each time point by variant. Shaded areas represent 95% confidence intervals. 1B-1E. Kaplan-Meier survival curves demonstrating time to negative PCR by viral variant (B) and vaccination status (D) and time to negative viral culture by viral variant (C) and vaccination status (E). Shaded areas indicate 95% confidence intervals from the survival curves. *P*-values represent log-rank testing comparing the sub-groups on each plot.

**Table 2.** Cox proportional hazards model of time to PCR conversion

| Covariate | Univariable Models |  | Multivariable Models |  |
| --- | --- | --- | --- | --- |
|  | Hazard Ratio (95%CI) | P-value | Hazard Ratio (95%CI) | P-value |
| Age (10 years) | 1.04 (0.87, 1.25) | 0.68 | 1.05 (0.87, 1.27) | 0.60 |
| Sex |  |  |  |  |
| Male | REF |  | REF |  |
| Female | 1.09 (0.60, 1.96) | 0.78 | 1.13 (0.62, 2.05) | 0.69 |
| Vaccination status |  |  |  |  |
| Unvaccinated | REF |  | REF |  |
| Vaccinated | 0.49 (0.23, 1.04) | 0.06 | 0.51 (0.23, 1.09) | 0.08 |
| Boosted | 1.00 (0.37, 2.71) | >0.99 | 1.16 (0.39, 3.45) | 0.80 |
| Variant |  |  |  |  |
| Delta | REF | 0.72 | REF |  |
| Omicron | 0.90 (0.50, 1.61) |  | 0.85 (0.45, 1.61) | 0.62 |

**Table 3.**
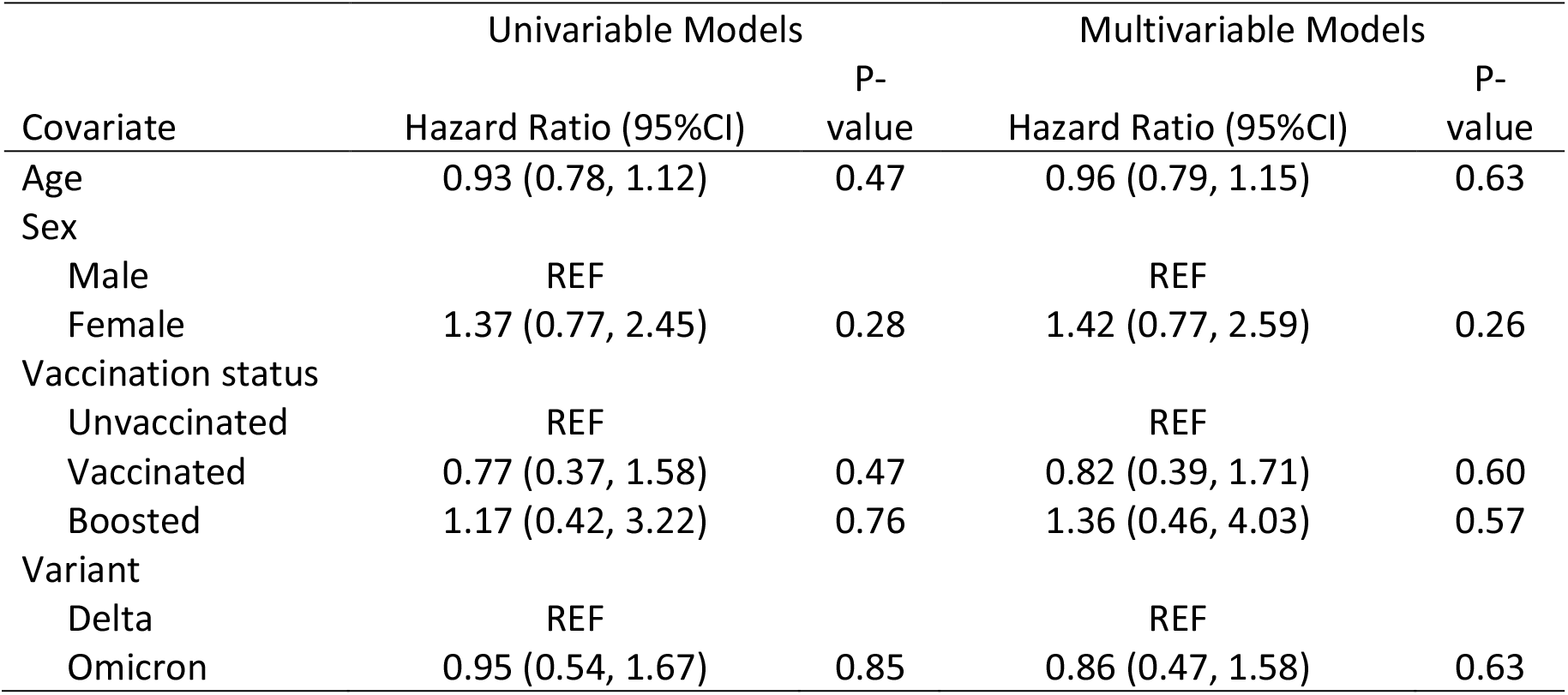
Cox proportional hazards model of time to culture conversion

**Figure 2.**
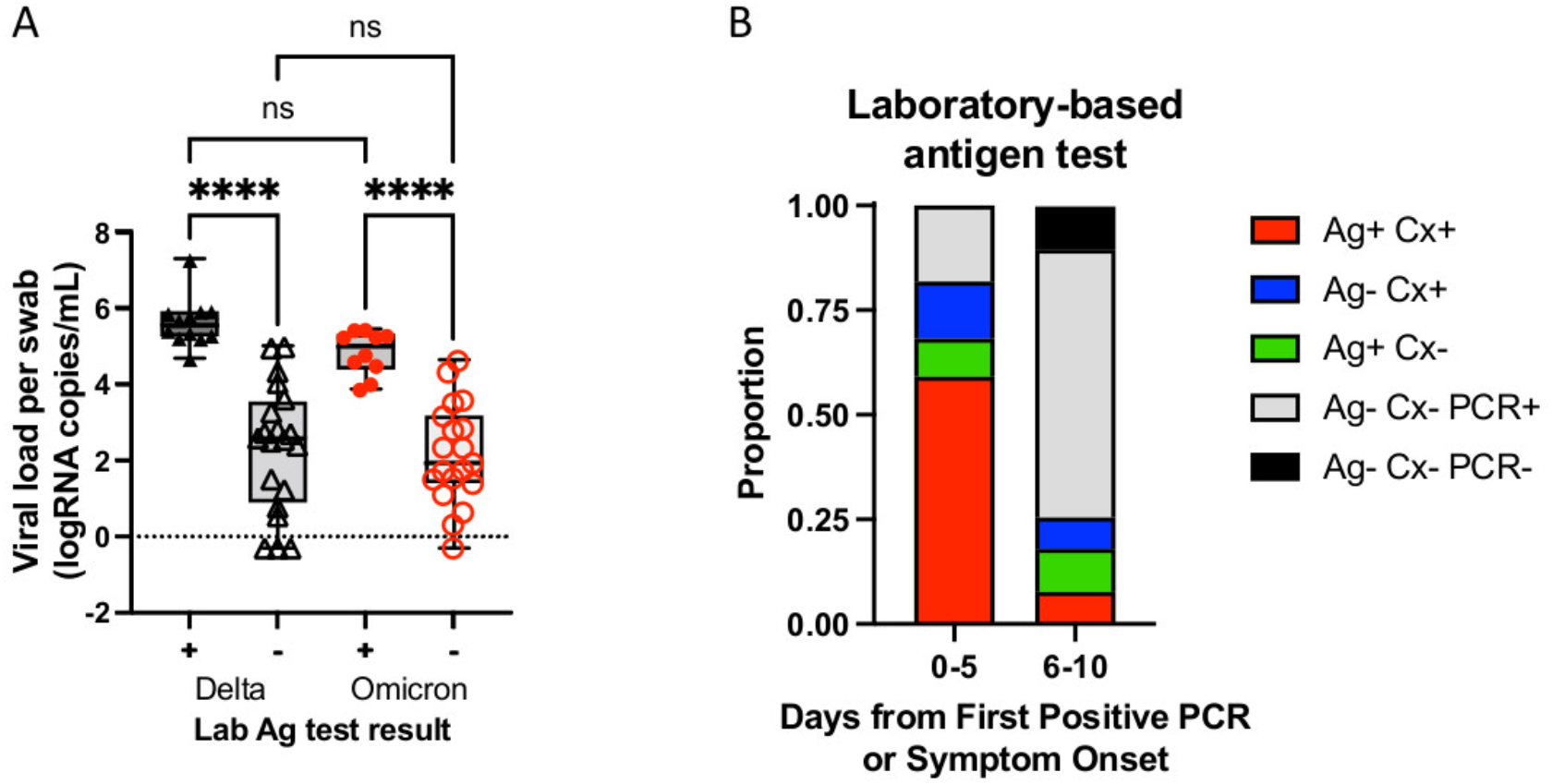
Antigen testing on samples in viral transport media simultaneously cultured. Samples (11 participants with delta infection and 11 participants with omicron infection) subjected to whole genome sequencing, viral load testing, and viral culture were simultaneously tested for antigen using the BinaxNow kit. (A) Viral load per swab for samples tested by laboratory-based antigen test results for delta and omicron infections. (B) Proportion of samples, delta and omicron infections combined, that were antigen test positive, culture positive and PCR positive (Ag+ Cx+), antigen test negative culture positive and PCR positive (Ag-Cx+), antigen test positive, culture negative and PCR positive (Ag+ Cx-), antigen test negative, culture negative and PCR positive (Ag-Cx-PCR+) or antigen negative, culture negative and PCR negative organized by days from first positive PCR or symptoms onset (0-5 or 6-10 days).

**Table 4.** Test validity of laboratory-based BinaxNow antigen testing compared to viral culture

| Sub-Cohort | Specimens |  | Sensitivity (95% CI) | Specificity (95% CI) |
| --- | --- | --- | --- | --- |
|  | Tested |  |  |  |
| Total | 61 |  | 72% (16/22), (50-88%) | 85% (33/39), (69-93%) |
| Delta | 33 |  | 69% (9/13), (38-89%) | 85% (17/20), (60-95%) |
| Omicron | 28 |  | 78% (7/9), (36-96%) | 84% (16/19), (59-95%) |
| Days 0-5 | 22 |  | 81% (13/16), (53-94%) | 67% (4/6), (18-95%) |
| Days 6-10 | 39 |  | 50% (3/3), (11-89%) | 88% (29/33), (71-96%) |

In this longitudinal cohort of individuals with symptomatic, non-severe COVID-19 infection, we found no difference in viral kinetics between omicron variant infection and delta variant infection or by prior vaccination history. Over 50% of individuals had replication competent, culturable virus at day 5, and 25% had culturable virus at day 8. Our cohort is limited to individuals with symptomatic, non-severe COVID-19 disease. A greater proportion of individuals infected with omicron had received their booster vaccine, although vaccination status was not associated with viral decay kinetics in multivariable models. Additional studies are needed to correlate viral culture positivity with confirmed transmission and to validate the utility of clinical antigen testing for defining optimal isolation periods.

## Methods

### Study Participants

Non-hospitalized individuals with positive SARS-CoV-2 PCR tests in the Mass General Brigham medical system were recruited. Adults over 18 years of age with a positive test in the medical health record were recruited, irrespective of indication for testing (i.e. for symptomatic disease, contact tracing, or work or pre-operative screening). For those who consented to participation, we conducted home visits three times weekly until negative PCR testing. At each visit, we obtained self-collected nasal swabs in viral transport media, which were transported to the laboratory within four hours of collection, aliquoted and frozen at -80°C until future testing. Symptoms, date of onset, and severity were recorded at each specimen collection. Symptomatic infections were defined as those with COVID-19-related symptoms at any point during the observation period.

### Viral load Quantification

Viral load quantification was carried out as previously reported^4^. Briefly, nasal swab fluids were centrifuged at 21,000 x g for 2 hours at 4°C to pellet virions. 750 μL TRIzol-LS™ Reagent (ThermoFisher) was then added to the pellets, and samples were subsequently incubated on ice for 10 minutes. 200 μL of chloroform (MilliporeSigma) was added to each sample, and the resulting mixtures were then vortexed and centrifuged at 21,000 x g for 15 minutes at 4°C. The clear aqueous layer was collected and combined with an equal volume of isopropanol (Sigma), 1.5 μL GlycoBlue™ Coprecipitant (ThermoFisher) and 100 μL 3M Sodium Acetate (Life Technologies); the resulting mixtures were briefly shaken and then incubated on dry ice. Samples were centrifuged at 21,000 x g for 45 minutes at 4°C to yield RNA pellets, which were washed with cold 70% ethanol before being resuspended in 50 μL DEPC-treated water (ThermoFisher). Using the US CDC 2019-nCoV_N1 primer and probe set (IDT) (https://www.cdc.gov/coronavirus/2019-ncov/lab/rt-pcr-panel-primer-probes.html) and N1 qPCR standards in 16-fold dilutions to generate standard curves, SARS-CoV-2 viral RNA was quantified. Each reaction consisted of extracted RNA, 1X TaqPath™ 1-Step RT-qPCR Master Mix, CG (ThermoFisher), forward and reverse primers, and the probe. Each sample was run in triplicate, and all plates contained two non-template control (NTC) wells. Positive and negative controls were run alongside all samples. To ensure appropriate sample quality, the Importin-8 (IPO8) housekeeping gene RNA level was quantified. The efficiency of the RNA extraction and qPCR amplification was assessed by quantifying the internal virion control RCAS ^5^ RNA level after spiking this viral mixture into each sample.

### SARS-CoV-2 culture

Viral culture was performed as previously reported in the BSL3 laboratory of the Ragon Institute of MGH, MIT, and Harvard^4^. Briefly, Vero-E6 cells (American Type Culture Collection) maintained in DMEM (Corning) supplemented with HEPES (Corning), 1X Penicillin 100IU/mL/Streptomycin 100 ug/mL (Corning), 1X Glutamine (Glutamax, ThermoFisher Scientific), and 10% Fetal Bovine serum (FBS) (Sigma) using Trypsin-EDTA (Fisher Scientific) were detached and seeded at 20,000 cells per well in 96w plates 16-20 hours before infection. Specimens were thawed on ice and filtered through a Spin-X 0.45um filter (Corning) at 10,000 x g for 5min. 25ul of the undiluted filtrate was added to four wells of a 96w plate and serial diluted (1:5) across half of the plate in media containing 5ug/mL of polybrene (Santa Cruz Biotechnology). Plates were centrifuged for 1 hour at 2000 x g at 37C. The positive control SARS-CoV-2 isolate USA-WA1/2020 strain (BEI Resources) was used in parallel for all assays. Plates were observed with a light microscope 7 days post-infection and documented wells with CPE. Supernatant of wells was harvested for RNA isolation using a QIAamp Viral RNA Mini kit (QIAGEN) for confirmation of the viral sequence.

### SARS-CoV-2 Whole Genome sequencing

Whole genome sequencing was performed as previously described^4^ following the Illumina COVIDSeq Test protocol. Libraries were constructed using the Illumina Nextera XT Library Prep Kit, then pooled and quantified using a Qubit High Sensitivity dsDNA kit (Invitrogen, Waltham, MA, USA). Genomic sequencing was performed on an Illumina NextSeq 2000, Illumina NextSeq 550, or Illumina NovaSeq SP instrument. Sequences with an assembly length greater than 24,000 base pairs were considered complete genomes, and those sequences were assigned a Pango lineage using the most up-to-date version of pangoLEARN assignment algorithm v2.4.2^6^. All sequences were deposited to GenBank and GISAID. The samples were submitted to NCBI with Bioproject Accession numbers PRJNA715749 or PRJNA759255.

### SARS-CoV-2 TaqPath RT-PCR Assay

Starting with Participant 200, samples were tested for spike gene target failure (SGTF) as an additional genotyping method of detecting Omicron cases following the TaqPath COVID-19 Combo Kit protocol (Thermo Fisher Scientific, Waltham, MA, USA). Nucleic acid was extracted using the MagMAX Viral/Pathogen II Nucleic Acid Isolation Kit on a Thermo KingFisher Flex purification system (Thermo Fisher Scientific, Waltham, MA, USA). Reverse transcription PCR (RT-PCR) was conducted on extracted samples using the Applied Biosystems 7500 Fast Dx Real-Time PCR Instrument (Applied Biosystems, Waltham, MA, USA), then analyzed for the presence of SARS-CoV-2 on ORF1ab, N gene, and S gene targets. SGTF was determined by amplification of SARS-CoV-2 for the ORF1ab and N gene targets with CT values <36 along with the lack of amplification for the S gene target.

### SARS-CoV-2 Spike gene amplification

Spike gene amplification was also performed as previously described^4^ to determine variant types for specimens with low viral load when whole genome sequencing was unsuccessful. cDNA synthesis was synthesized using Superscript IV reverse transcriptase (Invitrogen, Waltham, MA, USA) as per manufacturer’s protocols. cDNA amplification was performed using *in-house* designed primer sets that targeted codon 1-814 of the spike gene. PCR products were pooled for Illumina library construction using the Nextera XT Library Prep Kit (Illumina, San Diego, CA, USA). Raw sequence data was analyzed with PASeq v1.4 (https://www.paseq.org). Amino acid variants were identified at the codon level with perl code and the resulting variant file was used to determine SARS-CoV-2 variant type using Nextclade version 1.13.1 (https://doi.org/10.21105/joss.03773).

### Antigen testing using Abbott BinaxNow SARS-CoV-2 Rapid Antigen Assay

The AN VTM aliquots were thawed on ice and 50uL was transferred in a tube. The swabs from the BinaxNow kits were immersed into the liquid until it was fully absorbed^7^. The swabs were then tested according to manufacturer’s instructions. After 15min, for each test, a picture was taken, given a randomized ID and the results interpreted by three readers, blinded to the specimen ID. The outcome of each test was rated as positive, negative or discordant when not all three readers agreed.

### Statistical methods

We summarized demographic and clinical characteristics for individuals with delta and omicron variant infection and compared characteristics by sub-group with chi-squared testing for categorical variables and non-parametric testing for continuous variables. We graphically depicted viral decay by variant with a scatter plot and median of viral load over time since the first of symptom onset or index PCR test. To determine whether variant type or vaccination status was associated with virologic decay, we used the Kaplan-Meyer method to estimate the survivor function for two outcomes of interest: 1) time to conversion to negative PCR and 2) time to conversion to viral culture negative. For both outcomes we considered the earliest of date of symptom onset or first positive PCR test as the origin of observation. We defined the first day after the last positive PCR or positive culture as the date of exit. For individuals who had a positive PCR or culture on the final day of observation, they were censored as positive. For both outcomes, we constructed Kaplan-Meier curves of survival by variant and vaccination status.

We categorized vaccination status as unvaccinated, vaccinated, for those who had received two COVID-19 vaccinations (or a single dose of the Johnson & Johnson/Janssen vaccine) at least 14 days prior to enrollment, and boosted for those who had received three COVID-19 vaccinations (or a second dose of the Johnson & Johnson/Janssen vaccine) at least 14 days prior to enrollment. We compared time to PCR and culture version by sub-group using the log-rank method. We then fitted Cox proportional hazards models with both outcomes, and age, sex, vaccination status, and variant of infection as predictors. Finally, we graphed the distribution of viral load by BinaxNow antigen positivity and variant of infection and estimated the sensitivity and specificity of the tests compared to viral culture positivity, both overall and by time since symptom onset or first positive PCR test.

### Study approval

Study procedures were approved by the Human Subjects Institutional Review Board and the Institutional Biosafety Committee at Mass General Brigham. All participants gave verbal informed consent, as written consent was waived by the review committee based on the risk to benefit ratio of requiring in-person interactions for an observational study of COVID-19.

## Data Availability

All data produced in the present work are contained in the manuscript.

## Acknowledgments

The authors would like to thank study participants for willingness to engage in the study.

## Funding

This study was supported by the Massachusetts Consortium for Pathogen Readiness (grants to J.Z.L, J.E.L, M.B.G., M.J.S, and A.K.B.) and the Massachusetts General Hospital Department of Medicine (grant to J.M.V.). The BSL3 laboratory where viral culture work was performed is supported by the Harvard CFAR (P30 AI060354).

## Author contributions

J.B., M.G.B., J.M.V., J.Z.L, J.E.L, M.J.S, and A.K.B. designed the work. J.B., C.M., J.R., R.U., M.C.C., J.P.F., G.C., A.M.S., J.M., M.Y.L., A.S., T.L., A.K., M.M., Y.L., R.F.G., Z.R., S.L.I., G.C.C. and T.D.V. performed the work. J.B., J.R., R.U., M.C.C., J.P.F, Y.L., J.Z.L, J.E.L, M.J.S, and A.K.B. analyzed and reported the work.

## Notes

### Competing Interest Statement

The authors have declared no competing interest.

### Author Declarations

The Mass General Brigham IRB gave ethical approval for this work.

